# Burden of PCR-Confirmed SARS-CoV-2 Reinfection in the U.S. Veterans Administration, March 2020 – January 2022

**DOI:** 10.1101/2022.03.20.22272571

**Authors:** VA COVID-19 Observational Research Collaboratory

## Abstract

An essential precondition for successful “herd immunity” strategies for the control of SARS-CoV-2 is that reinfection with the virus be relatively rare. Some infection control, prioritization, and testing strategies for SARS-CoV-2 were designed on the premise of rare re-infection. The U.S. Veterans Health Administration (VHA) includes 171 medical centers and 1,112 outpatient sites of care, with widespread SARS-CoV-2 test availability. We used the VHA’s unified, longitudinal electronic health record to measure the frequency of re-infection with SARS-CoV-2 at least 90 days after initial diagnosis

We identified 308,051 initial cases of SARS-CoV-2 infection diagnosed in VHA between March 2020 and January 2022; 58,456 (19.0%) were associated with VHA hospitalizations. A second PCR-positive test occurred in 9,203 patients in VA at least 90-days after their first positive test in VHA; 1,562 (17.0%) were associated with VHA hospitalizations. An additional 189 cases were identified as PCR-positive a third time at least 90-days after their second PCR-positive infection in VHA; 49 (25.9%) were associated with VHA hospitalizations.

The absolute number of re-infections increased from less than 500 per month through November 2021, to over 4,000 per month in January 2022.

---

An essential precondition for successful “herd immunity” strategies for the control of SARS-CoV-2 is that reinfection with the virus be relatively rare. Some infection control, prioritization, and testing strategies for SARS-CoV-2 were designed on the premise of rare re-infection. Yet initial reports of re-infection have emerged.(1-5) National data are not currently available for the United States which link PCR-confirmed infections in the same individuals longitudinally in order to examine re-infection, report the total population burden of re-infection, and extend beyond 2021 and the emergence of the delta and omicron variants of concern.

The U.S. Veterans Health Administration (VHA) includes 171 medical centers and 1,112 outpatient sites of care, with widespread SARS-CoV-2 test availability. We used the VHA’s unified, longitudinal electronic health record to measure the frequency of re-infection with SARS-CoV-2 over time.

## METHODS

Patients with SARS-CoV-2 infection confirmed by PCR tests performed within VHA were identified in VA health care records between March 1, 2020 and January 31, 2022. All VA patients have a single unique record identification number. We identified reinfections as a positive PCR test occurring in VHA and at least 90-days after a prior positive PCR test also in VHA. We generated rates of reinfections per 1,000 cases per month and Poisson exact 95% confidence intervals for those rates because we were interested in the population burden of reinfections at each month. We identified associated hospitalizations as those in which patients were diagnosed during the 14 days preceding or 7 days following hospitalization, consistent with prior studies.(6)

## RESULTS

As shown in **Figure 1a**, the absolute number of re-infections increased from less than 500 per month through November 2021, to over 4,000 per month in January 2022. However, as shown in **Figure 1b**, the proportion of all PCR-confirmed cases in VHA in a given month that were reinfections was highest in spring of 2021 (exceeding 85 re-infections for each 1,000 infections in June 2021) and again climbed to approximately 55 re-infections for each 1000 new infection in January 2022.

**Figure 1:**
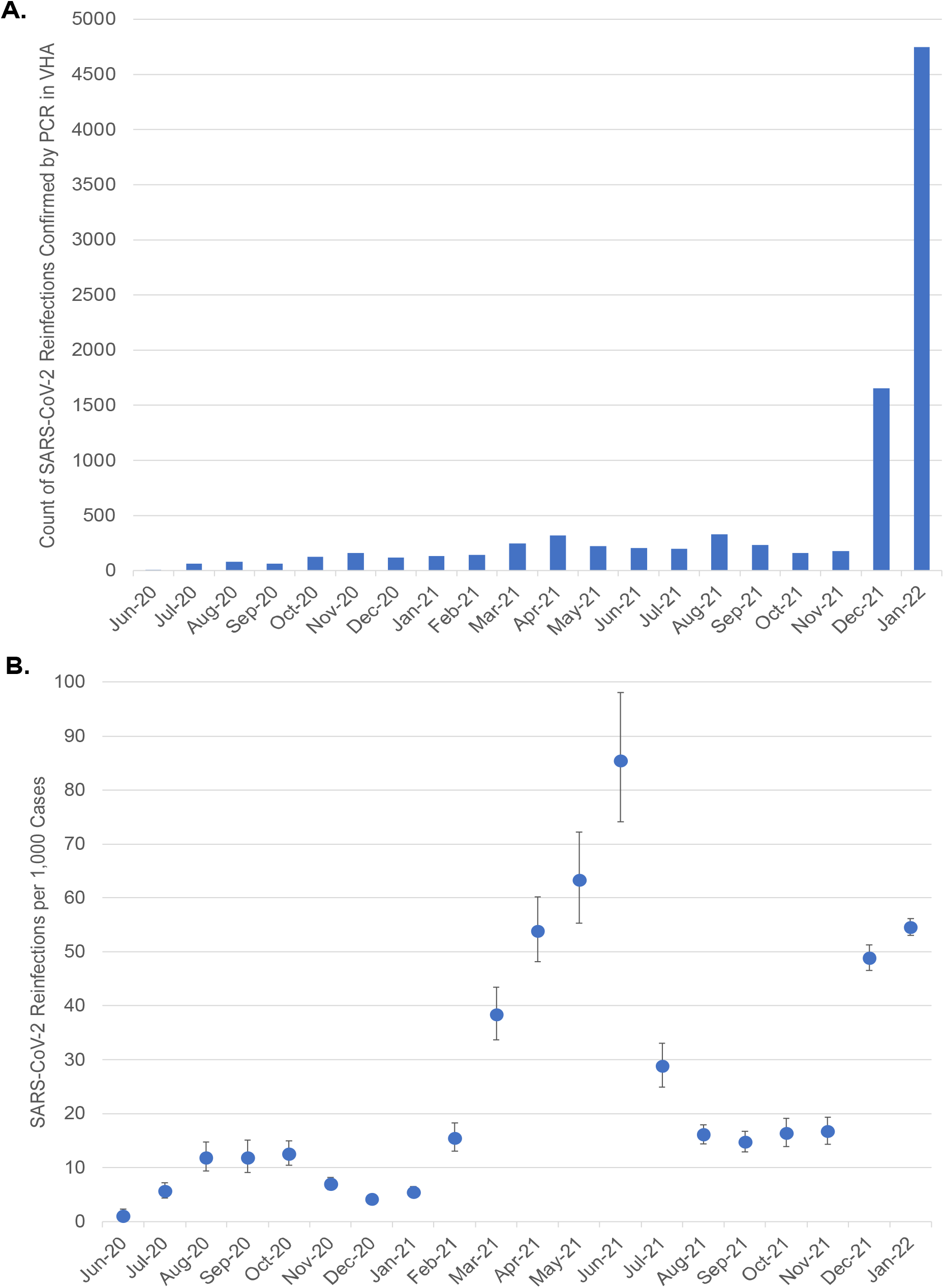
SARS-CoV-2 Reinfections confirmed by PCR in the Veterans Health Administration: (a) count of absolute number of re-infections; and (b) rate per 1,000 VHA SARS-CoV-2 infections identified in that month. Poisson exact confidence intervals for the rate are shown.

## CONCLUSION

These data suggest that re-infection with SARS-CoV-2 occurs regularly and can amount to a significant population level; these data are only a lower bound on the number of true re-infections, as they do not include infections that would be identified by testing done outside VHA or the untested. Re-infections as a proportion of all cases have not been limited to the period of the omicron wave. It is too soon to reliably measure the burden of disease among these patients. Given the rapid rise in absolute numbers, the absolute burden of SARS-CoV-2 re-infection may become substantial for health systems, caregivers, employers, and their potential infectivity to others. Testing and infection control strategies, systematic reporting, and clinical trial enrollment criteria that focus only on first confirmed SARS-CoV-2 infection risk will miss a large population of patients with the virus.

## Data Availability

All data are available in partnership with VA investigators via VA data approval processes.

## Acknowledgments

This work was supported by VA HSR&D C19-21-278 and C19-21-279. The Principal Investigators of the VA COVID-19 Observational Research Collaboratory are, in alphabetical order, Amy S.B. Bohnert PhD, C. Barrett Bowling MD, Edward J. Boyko MD, Denise M. Hynes PhD, George N. Ioannou MD, Theodore J Iwashyna MD PhD, Matthew L. Maciejewski PhD, Ann M. O’Hare MD, and Elizabeth M. Viglianti PhD. Troy Shahoumian led data analysis for this work. This report does not necessarily represent the views of the U.S. Government or Department of Veterans Affairs.

Please note that in all citations, the author of this should be only “VA COVID-19 Observational Research Collaboratory”. An individual named author appears in MedrXiv only because of MedrXiv’s requirements

## References

1. Cavanaugh AM, Spicer KB, Thoroughman D, Glick C, Winter K. Reduced Risk of Reinfection with SARS-CoV-2 After COVID-19 Vaccination - Kentucky, May-June 2021. MMWR Morb Mortal Wkly Rep. 2021;70(32):1081–3.

2. Hansen CH, Michlmayr D, Gubbels SM, Molbak K, Ethelberg S. Assessment of protection against reinfection with SARS-CoV-2 among 4 million PCR-tested individuals in Denmark in 2020: a population-level observational study. Lancet. 2021;397(10280):1204–12.

3. Lo Muzio L, Ambosino M, Lo Muzio E, Quadri MFA. SARS-CoV-2 Reinfection Is a New Challenge for the Effectiveness of Global Vaccination Campaign: A Systematic Review of Cases Reported in Literature. Int J Environ Res Public Health. 2021;18(20).

4. Mao Y, Wang W, Ma J, Wu S, Sun F. Reinfection rates among patients previously infected by SARS-CoV-2: systematic review and meta-analysis. Chin Med J (Engl). 2021;135(2):145–52.

5. Sheehan MM, Reddy AJ, Rothberg MB. Reinfection Rates Among Patients Who Previously Tested Positive for Coronavirus Disease 2019: A Retrospective Cohort Study. Clin Infect Dis. 2021;73(10):1882–6.

6. Donnelly JP, Wang XQ, Iwashyna TJ, Prescott HC. Readmission and Death After Initial Hospital Discharge Among Patients With COVID-19 in a Large Multihospital System. JAMA. 2021;325(3):304–6.

